# Best Practice Manufacturing and Quality Standards for Bacteriophage Therapy Products: Australian Consensus Statements

**DOI:** 10.64898/2026.08.26.26361487

**Authors:** K. Watts, R. CY. Lin, S. A. Lynch, J. Warning, J. J. Barr, N. L. Ben Zakour, JA Campbell, J. Chan, L. Collie, M. Hedges, B. Hudson, A. Irwin, A. Khatami, A. Kicic, D. Laucirica, C. Lauter, KM Ling, R. Ng, N. Pavuk, R. Rahmatullah, H. A. Sinclair, E. Tucker, S. Vreugde, M. Warner, Z. Velickovic, J. R. Iredell

**Author notes:** Corresponding author. Professor Jonathan R Iredell.

## Abstract

**Objective:** As antimicrobial resistance (AMR) continues to threaten global public health, bacteriophage therapy products (BTPs) offer a promising alternative to conventional antimicrobials.

However, translation into routine clinical practice requires best practice standards for manufacturing and quality control to ensure the consistent safety, quality, and reliability of personalised BTPs produced for individual patients or small cohorts.

**Design:** A modified Delphi methodology was used to develop consensus statements, engaging experts from Australia’s National Bacteriophage Therapy Regulatory Working Group across the fields of clinical microbiology, phage biology, good manufacturing practice (GMP), regulatory science, and government. The process comprised three iterative phases: (1) structured statement development, (2) an anonymous REDCap survey, and (3) a hybrid consensus meeting. The strength of evidence and recommendations was assessed using the GRADE (Grading of Recommendations Assessment, Development and Evaluation) framework.

**Results:** Consensus was reached on 35 statements to provide best practice manufacture and quality control guidance for BTPs. These statements address requirements for phage identification and characterisation; define the point at which GMP-aligned processes commence for ubiquitous phages; outline quality control expectations for phage active pharmaceutical ingredient (pAPI) production and maintenance of BTP and host cell repositories. Additional guidance covers quality management systems, including documentation, traceability, and governance.

**Conclusion:** These consensus statements provide comprehensive best practice recommendations for the manufacture and quality control of BTPs in Australia. By promoting consistent, safe, and quality-assured approaches to personalised BTPs, they aim to facilitate clinical implementation while remaining aligned with existing international pharmacopoeial standards and regulatory frameworks.

**Significance of this study:** *What is already known on this subject?:*

- Bacteriophage Therapy Products (BTPs) have re-emerged as a potential solution to antimicrobial resistance (AMR), supported by growing clinical evidence and global research activity. Data available to the regulatory working party includes more than 70 high-risk patients treated with dozens of different bacteriophages under a national open-label protocol (unpubl.).
- Published guidance from the European Pharmacopoeia, European Medicines Agency (EMA), UK Medicines & Healthcare products Regulatory Agency (MHRA), and World Health Oragnization (WHO) exists, yet no practical standards for personalised BTP manufacture have been established.

*What are the new findings?:*

- Recommendations have been made to define minimum quality and manufacturing stasndards for the safe, consistent production of BTPs.
- This is the first expert endorsed, risk-based framework to support the translation of personliased BTPs into routine clinical practice, cobering manufacture, repository management, quality control and supply.

*How might it impact on clinical practice in the foreseeable future?:*

- These consensus statements, developed by Australia’s National Bacteriophage Therapy Regulatory Working Group, can assist researchers, clinicians and regulatory authorities to develop BTP production protocols.

## Background

Antimicrobial resistance (AMR) is one of the top ten global public health threats and is projected to contribute to 169 million deaths between 2025-2050.^1^ The paucity of novel antimicrobial development to counter AMR has renewed interest in bacteriophage therapy products (BTPs) as a potential therapeutic alternative, ^2^ making BTP the most prevalent ‘non-traditional’ antimicrobial product globally. ^4^ Evidence from compassionate use programs demonstrates that BTPs are generally well tolerated, and therapeutically promising with 78.8% (n=1,500/1,904) of treated patients experiencing clinical improvement.^3^

While the World Health Organization (WHO) outlines a global roadmap for BTP implementation in their 2025 report, regulatory harmonisation remains challenging due to inconsistent classification of BTPs across jurisidctions (Table 2). For commercially developed BTPs, there are clear regulatory pathways applying to medicines and biological products. The challenge is the lack of standards for manufacturing and quality control of personalised BTP production, where products are selected, adapted, or prepared for an individual patient or small patient cohort.^5^ Despite progress to address this challenge by the European Pharmacopoeia chapter on phage therapy medicinal products, the European Medicines Agency (EMA) concept paper on the development and manufacture of human medicinal products specifically designed for phage therapy, and the UK Medicines & Healthcare products Regulatory Agency (MHRA) guidance on the therapeutic use of bacteriophages^6–8^, important practical questions remain regarding the selection and control of ubitiqous starting material, particularly given that bacteriophage are isolated from a wide range of natural sources.^9^ Key considerations include phage and host characterisation, traceability, repository governance, quality control, management of the transition between pre-Good Manufacturing Practice (GMP) to GMP activities, and the associated documentation requirements.

**Table 1 –.**
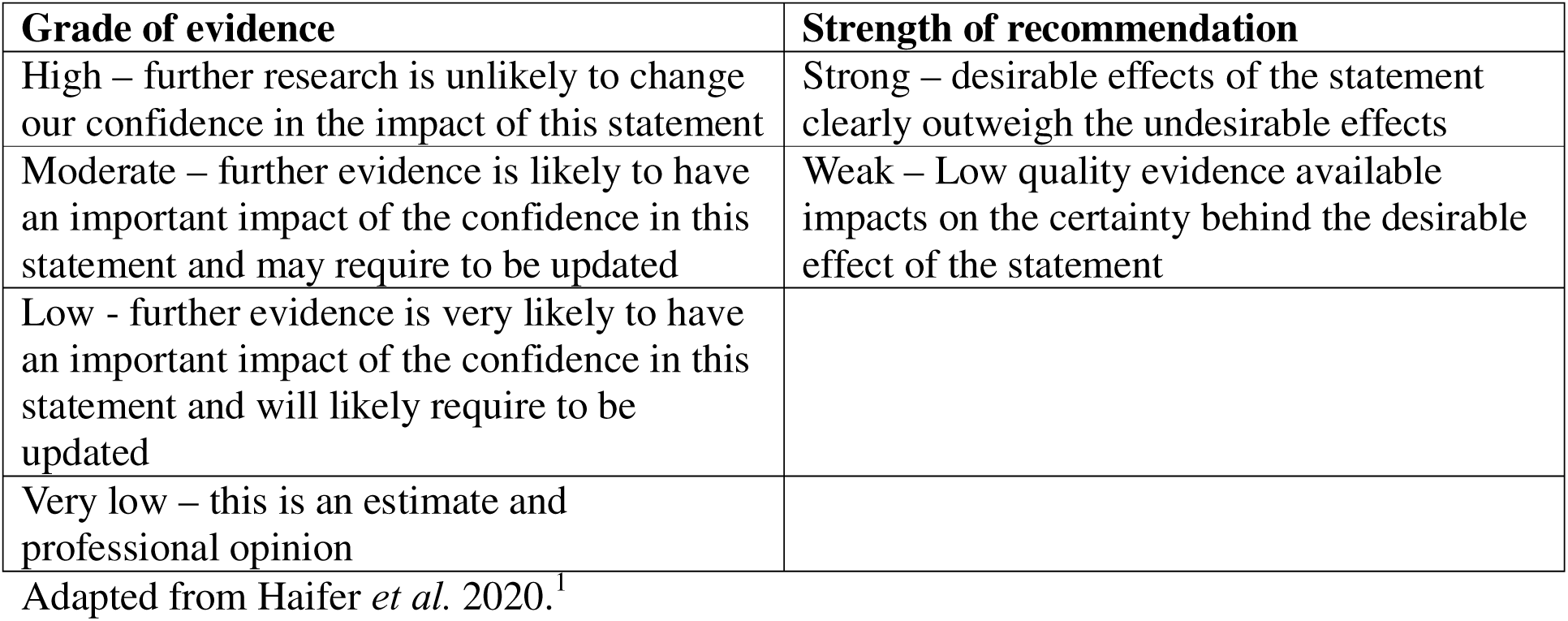
GRADE tool.

| Grade of evidence | Strength of recommendation |
| --- | --- |
| High – further research is unlikely to change our confidence in the impact of this statement | Strong – desirable effects of the statement clearly outweigh the undesirable effects |
| Moderate – further evidence is likely to have an important impact of the confidence in this statement and may require to be updated | Weak – Low quality evidence available impacts on the certainty behind the desirable effect of the statement |
| Low - further evidence is very likely to have an important impact of the confidence in this statement and will likely require to be updated |  |
| Very low – this is an estimate and professional opinion |  |

Adapted from Haifer *et al.* 2020.<sup>1</sup>

**Table 2 –.** Globally diverse regulatory approaches for BTP.

|  | Regulatory classification | Regulatory Pathway and Key Regulatory Features |
| --- | --- | --- |
| Belgium | Phage Therapy Medicinal Product | Magistral Preparations<sup>2</sup> |
| Estonia | Biological medicinal product | Compassionate use prepared at GMP standard in industry and exemption for hospital non-profit manufacture of personalized ATMPs<sup>3</sup> |
|  |  | The manufacturing of the final drug product should be GMP-compliant<sup>4</sup> |
| France | Medicinal products | Compassionate use regulated through the National Agency for the Safety of Medicines and Health Products (ANSM)<sup>5</sup> |
| Georgia | Pharmaceuticals | Magistral preparation in pharmacies specially licensed by the Georgian Ministry of Healthcare<sup>6</sup> |
| Germany | Medicinal products | Section 55 (8) AMG, all medicinal products must be manufactured in accordance with recognised pharmaceutical rules<sup>7</sup> |
| United Kingdom | Biological medicinal product | MHRA Regulatory considerations for therapeutic use of bacteriophages in the UK,<sup>8</sup> with structure framework under development |
|  |  | Compassionate use and GMP mandatory for domestic BTP production<sup>4,9</sup> |
|  |  | MHRA funding for engineered phage<sup>10</sup> |
| United States | Biological Products | Phage therapy medicinal products (PTMPs) manufacture must follow standards like GMP, preclinical research, and clinical trial<sup>11</sup> |
|  |  | FDA allows the extended access to the INDs of |
|  |  | phage therapy under exceptional situations for patients when they cannot enrol in clinical trials<sup>12</sup> |
| Canada | Biological drugs | Clinical trials under Food and Drug Regulations, Part C, Division 5:“Drugs for Clinical Trials Involving Human Subjects” GMP compliance is not required<sup>4</sup> |
| China | Innovative biological products | Phage products with fixed ingredients should be regulated as innovative biological products and personalized phage therapies need to go through IIT under Management Measures for Clinical Research<sup>13</sup> |
| Israel | Unclassified | Compassionate use |
| India | Unclassified | Compassionate use coordinated by the Central Drugs Standard Control Organization<sup>14</sup> |
| Poland | Unclassified | Compassionate use under European Medicines Agency only administered to patients in the Phage Therapy Unit<sup>15</sup> |
| Russia | Pharmaceutical | Magistral Preparation is forbidden<sup>14</sup><br>Only Mikrogen, the National manufacturer of immunobiological products in Russia, is authorized by the State Register of the Ministry of Health of the Russian Federation to manufacture market medicinal phage cocktails<sup>16</sup> |
| Australia | Medicinal products | Special Access Scheme (compassionate use) and clinical trials under Phage Australia uniquely adopts the Standardized Treatment and Monitoring Protocol for Adults and Pediatric Patients (STAMP) protocol to evaluate the clinical process for phage therapy<sup>17-19</sup><br>TGA approved 3-year GMP exemption for small batch bacteriophage therapy products while standards are developed<sup>20</sup> |

ATMPs - Advanced Therapy Medicinal Products; GMP - Good Manufacturing Practice; IND – Investigational New Drug; MHRA - Medicines and Healthcare products Regulatory Agency; TGA - Therapeutic Goods Administration; Magistral Preparation - allows a pharmacist to produce medicinal products based on a physician’s prescription for each patient in pharmaceutical standards

Australia’s National Bacteriophage Therapy Regulatory Working Group was established by the New South Wales Office for Health and Medical Research in February 2022 to provide expert input into the regulation of BTP in Australia, with membership from Phage Australia, taking BTP from the bench to bedside under their STAMP protocol, ^10^ and the Australian Therapeutic Goods Administration (TGA) and the Office of the Gene Technology Regulator (OGTR). The Regulatory Working Group sought to generate expert-informed consensus statements to guide the development of quality standards for BTP production in Australia.

The aim of the consensus process is not to replace existing international guidance, but to translate and extend these principles within the Australian context, providing a practical framework for the manufacture and supply of personalised phage therapies which aligns with existing manufacturing principles.

## Methodology

Given the absence of national guidance for BTP manufacturing, a Modified Delphi approach (Figure 1) to support structured expert consensus building was developed in accordance with the Accurate Consensus Reporting Document (ACCORD) guidelines. The methodology encompassed structured expert statement development, iterative data collection, and thematic synthesis, ^11^ with unanimous expert agreement without reservation.

**Figure 1 –.**
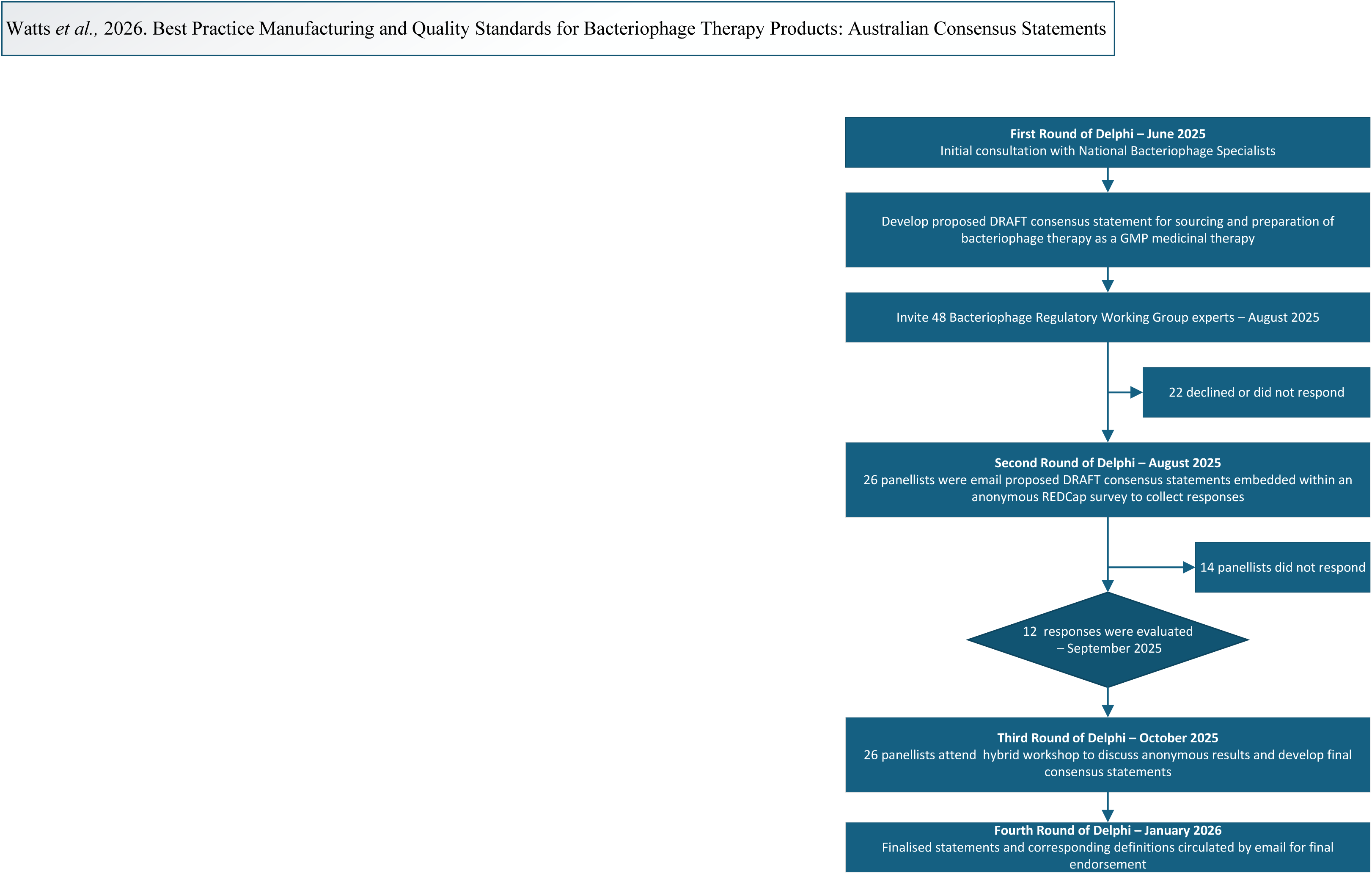
Modified Delphi Methodology.

All 48 members of Australia’s National Bacteriophage Therapy Regulatory Working Group were invited to participate in the study. In total, 26 experts spanning clinical microbiology, phage biology, good manufacturing practice (GMP), regulatory science, and government formed the Modified Delphi Working Group, constituting the panel for all subsequent rounds. The remaining members of the Regulatory Working Group are included in the acknowledgements.

### Round 1: Statement Development

A government regulatory and GMP expert undertook initial interviews with the expert bacteriophage researchers to develop an initial draft set of 43 consensus statements.

Statements were grounded in existing regulatory frameworks, pharmacopoeia standards, and applicable international guidance documents, encompassing topics including phage identification, repository requirements, GMP expectations, active pharmaceutical ingredient (API) production, and clinical quality controls. Each statement was applied a grade according to the level of evidence available and strength of recommendation as per the Grading of Recommendations Assessment, Development and Evaluation (GRADE) assessment (Table 1)^.12,13^

### Round 2: Anonymous Stakeholder Survey

The broader Delphi process was initiated through a structured, anonymous survey administered via REDCap. Participants were presented with each draft statement and asked to indicate acceptance or rejection; where statements were rejected, participants were invited to provide written recommendations for amendment. The inclusion of open-text responses permitted all participants to contribute freely without peer influence and facilitated identification of areas requiring clarification or substantive revision. This qualitative component is consistent with the ACCORD framework, which endorses the capture of expert reasoning rather than reliance on numerical agreement alone.^14^

In total, 12 responses were received (response rate: 67%). Of the 43 statements presented, 10 (23%) were accepted without modification. The remaining 33 statements were identified as requiring further deliberation and were carried forward to Round 3.

### Round 3: Hybrid Consensus Meeting

A full-day hybrid consensus meeting was convened in October 2025 at the New South Wales (NSW) Ministry of Health. Anonymised thematic summaries of Round 2 responses and corresponding proposed statement revisions were presented to the panel prior to deliberation. Each statement was discussed individually, with panellists providing expert insight, clarificatory context, and regulatory commentary, maintaining the iterative and structured principles of Delphi methodology. Proposed amendments were openly debated, and final statement wording was determined by unanimous agreement.

Through this iterative process, the total number of statements was consolidated from 43 to 35, reflecting consensus on removal of duplicating testing in the early phage identification and characterisation phase, refinement for precision and clarity, and removal of overlapping GMP content existing in published guidelines.

### Round 4: Finalisation and Endorsement

The finalised 35 consensus statements represent harmonised expert positions on the quality, safety, identification, manufacturing, repository management, and, regulatory considerations applicable to phage therapy products in Australia. Statements address foundational definitions (Statements 1–3); phage identification and characterisation requirements (Statements 4–6); repository and where GMP-aligned processes begin (Statements 7–12); active pharmaceutical ingredient (API) production and quality control (Statements 13–25); working phage API (pAPI) and bacterial cell repository maintenance (Statements 26-29); formulation and packaging (Statements 30-33); and documentation, traceability, and governance (Statements 34–35). To confirm consensus, in January 2026, endorsement was sought on the finalised statements and accompanying definitions from all Delphi members via email.

Phage Australia received research support and funding from NSW Health to further develop bacteriophage and to lead the National Bacteriophage Therapy Regulatory Working Group and the development of the consensus statements. Panel members were not remunerated for their contribution. Final consensus statements were endorsed by Phage Australia and the NSW Ministry of Health, reflecting the broad expertise and institutional agreement underpinning this work.

## Results

### Defining the production of BTPs

Statement 1: Phage preparations contain naturally occurring or genetically modified phage, independent of source and, may be used to treat or prevent human or veterinary bacterial infections.^7^ Grade: High, Strength: Strong.

Statement 2: Phage sourced from a third-party provider or legacy phage stock must meet the standards of both the local jurisdiction and requirements applicable for BTPs in Australia.

Grade: High, Strength: Strong.

Phage supplied by a third-party source must be accompanied by documentation regarding how the phage was propagated and a Certificate of Analysis (CoA) where feasible, to allow for an appropriate assessment of the quality of the phage to ensure safety for use.

Statement 3: Facilities preparing phage must be aware of and comply with relevant local laws and regulations. The design and construction of the manufacturing facility is suitable to the type of processing conducted, with separation of manufacturing areas for minimising laboratory error. Grade: High, Strength: Strong.

Phage preparation may involve manufacturing activities or pharmacy compounding and therefore requires access to appropriate governance and oversight arrangements. These may include a Drug and Therapeutics Committee (DTC) or Work Health and Safety (WH&S) Committee, and where genetically modified phage are used, an Institutional Biosafety Committee (IBC), constituted according to relevant legislations. A risk-based approach should be applied to determine how quality and safety requirements are met, recognising the inherent variability of biological starting materials, for example through segregation of phage manufacturing activities to minimise contamination risk.

### Phage Identification

Research-level characterisation assays, including identity, host range, genomic sequencing and, exclusion of undesired genes, are essential for phage selection but do not in and of themselves satisfy GMP requirements.^15^ Facilities conducting, or intending to conduct, studies to support regulatory submissions must therefore comply with Good Laboratory Practice (GLP) to ensure the reliability, integrity, and regulatory acceptability of phage safety data.^16^

Statement 4: Phage must be propagated using qualified equipment and reagents that must not adversely affect the quality or safety of the products. Grade: High, Strength: Strong.

All materials used during the propagation of phage, including culture media, buffers, consumables, and laboratory ware, must meet defined quality standards, for example ISO 90001 for laboratory quality management and ISO 17025 for testing laboratories, to minimise the risk of introducing contaminants such as adventitious viruses and endotoxins.

Animal-derived materials may be used during the early phases of phage identification and preparation. Where applicable, all reagents must be supported by manufacturer documentation, including Certificates of Analysis (CoA), to enable appropriate quality assessment and support the satisfactory quality control of the product.

Statement 5: Phage must be characterised and annotated according to genotypic markers, with a specific search for bacterial virulence-enhancing markers, integration (lysogeny) markers and antimicrobial resistance genes they may encode. Grade: Moderate, Strength: Strong.

Genomic characterisation or validated alternative characterisation strategy should be used to confirm the viability and purity of the phage^17^ so as to facilitate selection of the appropriate phage-host-patient combination and intended therapeutic indication.^18^

Statement 6: Isolation of each single phage seed lot is tested for its lytic activity to determine the receptive bacterial host. Grade: High, Strength: Strong.

Phage for therapeutic use are expected to be obligately lytic. Where lysogeny is proposed as a critical component of the mechanism of action, sufficient evidence must be provided to demonstrate no unacceptable risk to patient safety.^8^

To prevent the unwanted drift of properties in subcultures, the production of medicines obtained by microbial culture is best based on a validated system of master source and working repositories.^19,20^

### Master source repository (phage seed and bacterial cell)

Statement 7: Phage must be stored in a dedicated phage repository with an acceptable Quality Management System (QMS). Grade: High, Strength: Strong.

The master source repository relates to the storage and maintenance of raw biological materials (e.g., phage seed lots and bacterial cells) and is not required to meet GMP standards. These activities must, however, conform with Good Laboratory Practice (GLP). Storage may be performed by a third-party organisation, such as a research laboratory, provided it operates within a validated QMS e.g., ISO 9001. This master source repository is distinct from laboratory research in that materials are retained for future clinical or manufacturing use, rather than exploratory experimentation, and therefore require a higher level of governance, documentation, and quality oversight.^7^

Statement 8: Bacterial cells intended to propagate phage must be stored in a dedicated repository with an acceptable QMS. Grade: High, Strength: Strong.

Bacterial host cells used for phage active pharmaceutical ingredient (pAPI) production must be derived from a well characterised bacterial master repository.^7^ The bacterial cell repository must include identification of bacterial cell using a suitable method, microbial purity, viability, phage sensitivity and absence of detrimental phage particles that may affect the quality of pAPI is confirmed.

This master source repository ensures consistency, traceability, and regulatory compliance of starting materials for pAPI production, in accordance with an approved monograph or defined production framework.^20^

### Good Manufacturing Practice requirements

Application of PIC/S PE009 Guide to good manufacturing practice for medicinal products,^21^ or PE010 Guide to good practices for the preparation of medicinal products in healthcare establishments,^22^ begins once the identified and characterised phage has been extracted from the host and classified as the pAPI starting material.

Statement 9: Phage production facilities must have and comply with a QMS that meets the requirements of GMP principles, and to comply with existing requirements for clinical trials. Grade: High, Strength: Strong.

GMP represents the minimum standard that must be met by manufacturers of phage for clinical use. Adherence to GMP principles ensures that production processes consistently deliver products of appropriate quality, are fit for their intended use, and comply with approved product specification.

The QMS forms the foundation of an organisation’s documented policies, procedures, and governance framework. It is a multifunctional system that ensures operational processes required for product or service provision are established, monitored, audited, and continuously improved. A robust QMS supports risk management, regulatory compliance, and quality oversight, while also contributing to improved leadership, customer experience, employee engagement, and overall organisational effectiveness.^23^

Statement 10: Facilities and equipment must be located, designed, constructed, adapted and maintained to ensure operational compliance with GMP principles, including separation of areas for minimising laboratory error/contamination, and to comply with existing requirements for clinical trials. Grade: High, Strength: Strong.

When preparing pAPI for clinical use, the manufacturing process requires a specialised facility where phage propagation, prior to filtration, be performed in a minimum EU GMP Grade C cleanroom, equivalent to ISO 14644 Class 7,^25^ and that fill and finish be performed in a minimum EU GMP Grade A negative pressure Biological Safety Cabinet (BSC) housed in a EU GMP Grade B cleanroom or equivalent.^26^

Statement 11: The policies and standard operating procedures (SOPs) used to prepare phage must be clearly defined, documented and regularly reviewed in a systematic way to ensure phage are safe for use, prepared or manufactured consistently, and comply with the specifications for the products. Grade: High, Strength: Strong.^17^

Statement 12: All facilities must have an adequate number of trained personnel with the necessary qualifications and practical experience to prepare/manufacture phage for clinical use, including practical skills and quality control. Grade: High, Strength: Strong.

Clearly defined procedures, regular review of operating practices, and appropriately trained personnel are fundamental to ensuring that phage products are prepared consistently, meet predefined quality specifications, and are safe for clinical use. These requirements should be embedded as core elements of the QMS across document control, training and competency management, quality assurance, and continuous improvement. SOPs should be formally approved, version-controlled, and periodically reviewed, while personnel qualifications, training records, and ongoing competency assessments should be maintained and monitored.

### Phage Active Pharmaceutical Ingredient production

In pAPI preparation, impurities such as endotoxins must be maintained below the acceptable limits, according to the application, whether topical, intravenous or intrathecal, with product quality assessed through evaluation of stability, sterility, and pH to support safe clinical use.^17,27^ Upstream process development enables manufacturing scale preparation focusing on optimisation of key culture parameters, including bacterial density, multiplicity of infection, and culture medium to refine production.^28,29^

Statement 13: Phage must be propagated using qualified equipment and reagents that must not adversely affect the quality or safety of the products. Animal-derived material must be appropriately qualified and removed later in the processing. Grade: High, Strength: Strong.

All materials used during the propagation of phage such as media, buffers and labware, must meet defined quality standards. Non-animal-based inputs are recommended where possible to reduce bioethical concerns, zoonotic risk, and a simplified regulatory pathway. Non-animal reagents are not required to comply with stringent safety testing required for animal-origin materials e.g., foetal bovine serum.^30^ All reagents used require documentation by each manufacturer, i.e., the CoA, to support the satisfactory quality control of the product.

Statement 14: Phage amplification, expansion and extraction must adhere to GMP standards, validated SOPs and equipment calibration. These processes can be achieved through open or closed systems, dependent upon the risk assessment, to ensure product safety and quality.

Grade: High, Strength: Strong.

Phage are amplified using a susceptible bacterial host by infecting the culture, allowing phage replication within the bacteria, and harvesting the newly produced phage following bacterial lysis, resulting in a high-titre preparation for downstream purification.

Amplification may be performed using closed or open systems, subject to a risk-based assessment. Closed systems minimise aerosolisation, reduce contamination risk, and limit occupational exposure. Open systems, involving exposure of the product to the surrounding environment, may be used where justified, provided they are conducted within appropriate containment, gowning, and environmental controls. Final fill and finish operations should be performed in an EU GMP Grade A environment to ensure both product sterility and personnel safety, with risk assessments considering contamination control, aerosol generation, and operator exposure in accordance with PIC/S PE009^21^ and EU GMP Annex 1 principles.

Statement 15: pAPI must be purified using qualified equipment and reagents that do not adversely affect the quality or safety of the products. Heavy metals or organic solvents, must be appropriately qualified and shown to be removed later in the processing. Grade: Moderate, Strength: Strong.

Residual amounts of heavy metals and organic solvents may pose toxicological risks to patients, affect stability, and create batch-to-batch variability. Their removal is necessary to ensure product quality, safety, and compliance with regulatory standards. The downstream purification process is essential in reducing impurities derived from the production process.^29^

Statement 16: DNA and DNA fragments derived from host bacterial cells are acceptable at < 10 nanograms (ng) of residual host cell DNA per therapeutic dose Grade: High, Strength: Moderate.

The World Health Organisation (WHO) recommend a maximum of 10 ng of human host DNA per therapeutic dose, and this is increasingly applied as a risk-based benchmark in GMP-aligned phage manufacturing.^31,32^

Statement 17: Purity and quality of pAPI should be validated phenotypically through laboratory assessments and genomically through Next-Generation Sequencing (NGS), or the most appropriate test method. Grade: High, Strength: Weak.

For safety and consistency, regulatory bodies expect full compliance to ICH Q6B specifications for phage characterisation, purity and safety testing of biologics derived from live organisms.^33,34^ Phenomic characterisation would include morphology (head and tail structure), determination of structural protein, host range analysis and resistance, virulence (lytic or lysogenic potential), thermal and acid-base stability,^8,35,36^ while genomic characterisation would include sequencing to identify whether phage carry genes that could enhance the virulence of their bacterial hosts.

Statement 18: Phage must undergo qualified validated methods that provide evidence of *in-vitro* efficacy against a defined bacterial strain prior to inclusion in the Working Phage Repository. Grade: High, Strength: Strong.

Statement 19: Phage pAPI must undergo suitable bioburden reduction into validated sterile final packaging. Grade: High, Strength: Strong.

pAPI are living organisms, unable to be terminally sterilised. To adequately reduce the risk of microbial (bacterial) contamination, filtration of the pAPI can be achieved using a validated and integrity tested 0·22-micron filtration, or the most appropriate method, in accordance with PIC/S PE009.^21,37,38^ See also Statements 12, 16.

### pAPI Quality Control

Quality Control (QC) is fundamental to ensure the identity, purity, and quality of each phage product for use in clinical practice. QC of phage typically utilises a series of characterisations assays prior to direct QC release tests before a phage product is administered to a patient.^28^

The TGA recognises the limitations of terminal sterilisation for phage and biologicals,^37^ where the PIC/S Annex 1 explicitly states that aseptic processing is acceptable when terminal sterilisation is not feasible for biologics.^25^

Statement 20: The manufacturer must perform a validated test for particulate contamination. Grade: High, Strength: Strong.

Unintended particulate matter in parenteral preparations consists of mobile undissolved substances, other than gas bubbles, which may originate from various sources. The level of particulate matter must be minimised and controlled, independent of its type.^39^

Statement 21: The manufacturer must ensure that pAPI undergoes endotoxin testing, with acceptable endotoxin limits defined according to the intended route of administration (Table 3). Consideration must be given to other available virulent toxin testing. Grade: High, Strength: Strong.

**Table 3 –.** BPT Endotoxin limits.

| Route of administration | EU limit |
| --- | --- |
| Intravenous | 5 EU/kg/hour |
| Intramuscular | 5 EU/kg/hour or 0.5 EU/mL |
| Intrathecal | 0.2 EU/kg/hour |
| Subcutaneous | 0.5 EU/mL |
| Inhaled | 5 EU/kg/hour |
| Topical | 100 EU/m<sup>2</sup> |

References USP <85> Bacterial Endotoxin Test,<sup>21</sup> and British Pharmacopoeia.<sup>22</sup>

Bacterial endotoxins, derived from Gram-negative bacteria, are the most common cause of pyrogenic reactions in pharmaceutical products. Medicinal products intended for parenteral administration must therefore be sterile and comply with bacterial endotoxin testing (BET), using pharmacopeial or otherwise validated methods.^40^ Acceptable endotoxin limits vary by route of administration and are defined in pharmacopeial guidance (Table 3), with more stringent limits applied to high-risk routes such as intrathecal administration.^41,42^ See also Statement 12.

In addition, manufacturers are required to assess and control the risk of N-nitrosamines, which are probable human carcinogens, in parenteral products, in accordance with European Pharmacopoeia general monographs and supporting guidance.^43^ General chapter 2·5·42 N- Nitrosamines in active substances is available to assist manufacturers.^44^

BTPs not intended to be sterile, such as oral or topical formulations, are not subject to specific endotoxin or N-nitrosamine limits but must comply with microbiological quality requirements for non-sterile products. Liquid preparations for nebulisation must be manufactured as sterile product.^45,46^

Statement 22: The manufacturer must have a validated and calibrated sterility test completed by a National Association of Testing Authorities (NATA) accredited authority. Grade: High, Strength: Strong.

TGA mandates that medicinal products intended for human use, especially those administered intravenously, meet strict sterility criteria. Certification obtained from a NATA accredited organisation confirms that the testing facility adheres to ISO 17025 standards, or the most appropriate standard, providing confidence in the reliability and reproducibility of results.

This is critical for regulatory submissions, batch release, and patient safety.

Statement 23: The manufacturer must ensure that pAPI intended for intravenous administration has a pH within the range of 4–9. Grade: High, Strength: Strong.

TGA expects intravenous products to maintain isotonicity and a physiologically compatible pH, supported by validated stability and compatibility data. Phage formulations that fall outside this pH range may cause irritation, haemolysis, or vascular damage upon infusion. Extremes of pH can also compromise the stability of phage medicinal products by degrading capsid proteins or reducing infectivity.

Statement 24: The Manufacturer must issue a CoA for the pAPI. Grade: High, Strength: Strong.

A CoA is required after final quality control testing to confirm that the phage medicinal product meets predefined specifications, as reported in these consensus statements, and is suitable for clinical or therapeutic use.

Statement 25: The manufacturers must have and comply with procedures for the cleaning and decontamination of facilities and equipment, to avoid any cross-contamination during the development or manufacturing steps. Grade: High, Strength: Strong.

Given the high titres of phage obtained *in vitro*, it is essential to establish validated cleaning and decontamination procedures to prevent cross-contamination during development or manufacturing.^28^ ISO 10705-3 provide structured frameworks for validating cleaning, including specific requirements for phage inactivation.^47^

### Working pAPI and bacterial cell Repository maintenance

Statement 26: The pAPI repository must comply with PIC/S PE009 requirements and be operated by an organisation accredited by a NATA-endorsed body to ISO 15189, with appropriate compliance certification in place. Grade: High, Strength: Strong.

Statement 27: The storage of the bacterial cell repository must comply with PIC/S PE009 requirements and be operated by an organisation accredited by a NATA-endorsed body to ISO 15189, with appropriate compliance certification in place. Grade: High, Strength: Strong.

ISO 15189 sets out the requirements for quality and competence to TGA manufacturing guidelines, including validated traceable processes for phage manufacture, ensuring that analytical methods used to characterise and release phage, meet internationally recognised standards, supporting downstream manufacturing and clinical use.^48^

Statement 28: The pAPI and bacterial cell repository maintenance and record keeping must be in line with PIC/S PE009 Part II Section 18. Grade: High, Strength: Strong.

Statement 29: The manufacturer must provide justification for the application of an expiry date and must consider stability testing as required. Grade: High, Strength: Strong.

Over time, the titre of purified phage preparations may decline due to environmental stressors, formulation variables or storage conditions. Conducting rigorous stability assays, including accelerated and real-time testing, enables determination of appropriate expiry dates and helps safeguard against under-dosing.

### Formulation and Packaging

Statement 30: The manufacturer must label the pAPI to identify the titre of viable PFU. It is recommended that the final pAPI contain a minimum titre of 10^9^ PFU/mL. ^49–51^ Grade: Moderate, Strength: Strong.

Statement 31: The formulation and packaging of pAPI must be appropriate for the intended route of administration^8^ and fit for their intended purpose. Grade: High, Strength: Strong.

Manufacturers must use sterile components (vials, stoppers, diluents) prior to final packaging, with packaging systems validated to minimise contamination risk during use. In addition, some plastics are incompatible with phage, as hydrophobic capsid surfaces may adsorb to materials such as polypropylene or polystyrene, reducing free phage titre, potentially affecting dose accuracy and efficacy. Selection of low-binding, phage-compatible materials (e.g. siliconised glass or medical grade polymers) is needed, with ongoing monitoring of potential leachable packaging during storage.^52^

Statement 32: Medicinal Phage must be transported under validated and monitored conditions. Grade: High, Strength: Strong.

Some phages are susceptible to inactivation by heat,^53^ light exposure,^54^ and physical shearing.^55,56^ Validation studies must therefore be conducted to validate stability of phage preparations during transportation and storage.

Statement 33: Transport packaging must comply with biological product shipping standards, but phages are exempt from UN3373 requirements. Grade: High, Strength: Strong.

Under UN3373 exemption 3·6·2·2·3·2, substances containing microorganisms that are non-pathogenic to humans or animals are not subject to these Regulations unless they meet the criteria for inclusion in another class.^57^ Phage are therefore exempt from UN3373 labelling as “live biological agent”, as they fall under IATA Category B.

### Documentation

Statement 34: Procedures must be in place to ensure the traceability of phage from source through to manufactured BTP. Grade: High, Strength: Strong.

Documentation enables the investigation of deviations or product quality failures, the implementation of corrective actions, and regulatory review. It also helps document product identity, consistency, and patient safety as phages can vary significantly depending on their source and processing history.

Statement 35: A system must be established and maintained to: (a) handle complaints made or concerns raised by any person in relation to the manufacture of phage products; and (b) ensure process and quality improvement functions and activities are carried out regularly Grade: High, Strength: Strong.

The process ensures that concerns regarding safety, quality, or compliance are addressed in a structured and timely manner. It helps in detecting recurring issues, systemic weaknesses, or opportunities for process improvements. Additionally, it reduces the risk of repeated failures by converting complaints and observations into preventive actions.

## Conclusions

In the context of escalating AMR, these best practice consensus statements deliver urgently needed guidance on acceptable standards for the safe and consistent quality of BTPs.

Throughout this process, the expert panel considered clinical, regulatory, process, and diversity-related dimensions to ensure that the resulting framework is both rigorous and implementable. The expert panel reached consensus that the collection, identification, and master repository storage of bacteriophages fall outside GMP requirements; however, all materials used during propagation must still meet defined quality standards. Under the relevant sections of PIC/S PE009 or PE010, GMP obligations apply once the characterised phage is extracted from the host and designated as the pAPI starting material. These consensus outcomes strengthen governance, quality assurance and reproducibility by providing a practical framework to support the safe integration of bacteriophage therapy into clinical trials and healthcare systems, enabling more timely access to a potentially lifesaving complementary intervention against AMR.

## Limitations

Although the Modified Delphi methodology relies on expert consensus rather than clinical outcome data, it is well suited to areas with limited empirical evidence and evolving regulatory frameworks and was necessary to address an urgent need for pragmatic guidance in a rapidly developing field. The expert panel was relatively small, comprising 26 panellists and a 67% response rate in Round 2; however, this level of participation is consistent with Delphi studies in highly specialised domains. While the statements were developed in close collaboration with the TGA to ensure regulatory relevance and feasibility, the panel was limited to Australian experts, which may affect generalisability to other jurisdictions.

Nevertheless, the co-design with regulators confers robustness, credibility, and immediate applicability within the Australian regulatory context.

## Contributors

Watts K: Conceptualisation; Investigation; Methodology; Project administration; Writing – original draft; Writing – review & editing.

Lin R: Conceptualisation; Investigation; Supervision; Validation; Writing – original draft; Writing – review & editing.

Lynch S: Investigation; Writing – review & editing.

Warning J: Conceptualisation; Investigation; Methodology; Writing – original draft; Writing – review & editing.

Barr JJ: Investigation; Writing – review & editing.

Ben Zakour N: Investigation.

Campbell JA: Investigation; Writing – review & editing.

Chan J: Investigation.

Collie L: Conceptualisation; Investigation; Writing – review & editing.

Hudson B: Investigation; Writing – review & editing.

Irwin A: Investigation; Writing – review & editing.

Khatami A: Investigation; Writing – review & editing.

Kicic A: Investigation; Writing – review & editing.

Laucirica D: Investigation; Writing – review & editing.

Lauter C: Investigation.

Ling KM: Investigation; Writing – review & editing.

Ng R: Investigation; Writing – review & editing.

Pavuk N: Investigation; Writing – review & editing.

Rahmatullah R: Investigation.

Sinclair H: Investigation; Writing – review & editing.

Tucker E: Investigation; Writing – review & editing.

Vreugde S: Investigation; Writing – review & editing.

Warner M: Investigation; Writing – review & editing.

Velickovic Z: Investigation; Supervision; Validation; Writing – review & editing.

Iredell J: Conceptualisation; Investigation; Supervision; Validation; Writing – review & editing.

All authors contributed to the investigation and interpretation of the consensus process, had access to the underlying data supporting the statements, and approved the final version of the manuscript.

## Data Availability

All data produced in the present work are contained in the manuscript

## Acknowledgements

We thank all current members of the Australian Bacteriophage Therapy Regulatory Working Group for their efforts to support the regulation of phage therapy in Australia, including: Helene Lebhar, Stephen Stick, Anton Peleg, Steven Tong, Vanessa Fitzgerald, Bruce Battye, Judith Mackson, Tony Gill, Jenny Hantzinikolas, Glenn Smith, Ting Lu, Alyce Maksoud, Avril Fortuin, Heidi Mitchell, Matt Davis, O’Mezie Ekwudu, Australian Pesticides and Veterinary Medicines Authority: Maria Trainer, Mariette Van den Berg, Dharma Purushothamam. Phage Australia team: Clare Fisher, Claire Elek. The Kids Research Institute Australia: Mitchell Hedges. Therapeutic Good Administration: Tony Manderson and Anna Hart. Office for Gene Technology Regulator: Cecily Goulart.

## References

1. Collaborators GBDAR. Global burden of bacterial antimicrobial resistance 1990-2021: a systematic analysis with forecasts to 2050. Lancet 2024; 404(10459): 1199–226.

2. Organisation WH. International Clinical Trials Registry Platform (ICTRP). 2025.

3. Uyttebroek S, Chen B, Onsea J, et al. Safety and efficacy of phage therapy in difficult-to-treat infections: a systematic review. Lancet Infect Dis 2022; 22(8): e208–e20.

4. Organisation WH. 2025 antibacterial agents in clinical and preclinical development: an overview and analysis. Geneva, ISBN 978-92-4-011309-1: World Health Organisation, 2025.

5. Fukaya-Shiba A, Ogata A, Kuribayashi R, et al. Regulatory considerations for developing phage therapy medicinal products for the treatment of antimicrobial resistant bacterial infections. Front Pharmacol 2025; 16: 1713471.

6. Use CfMPfH. Concept paper on the establishment of a guideline on the development and manufacture of human medicinal products specifically designed for phage therapy. Amsterdam: European Medicines Agency, 2023.

7. HealthCare EDftQoM. Phage therapy medicinal products. European Pharmacopoeia 116. 11.6 ed. Strasbourg: EDQM; 2026.

8. Agency MaHpR. Regulatory considerations for therapeutic use of bacteriophages in the UK. London: MHRA, 2025.

9. Iszatt JJ, Larcombe AN, Garratt LW, et al. Genome Sequence of a Lytic Staphylococcus aureus Bacteriophage Isolated from Breast Milk. Microbiol Resour Announc 2022; 11(12): e0095322.

10. Khatami A, Foley DA, Warner MS, et al. Standardised treatment and monitoring protocol to assess safety and tolerability of bacteriophage therapy for adult and paediatric patients (STAMP study): protocol for an open-label, single-arm trial. BMJ Open 2022; 12(12): e065401.

11. Logullo P, van Zuuren EJ, Winchester CC, et al. ACcurate COnsensus Reporting Document (ACCORD) explanation and elaboration: Guidance and examples to support reporting consensus methods. PLoS Med 2024; 21(5): e1004390.

12. Guyatt GH, Oxman AD, Vist GE, et al. GRADE: an emerging consensus on rating quality of evidence and strength of recommendations. BMJ 2008; 336(7650): 924–6.

13. Haifer C, Kelly CR, Paramsothy S, et al. Australian consensus statements for the regulation, production and use of faecal microbiota transplantation in clinical practice. Gut 2020; 69(5): 801–10.

14. Nasa P, Jain R, Juneja D. Delphi methodology in healthcare research: How to decide its appropriateness. World J Methodol 2021; 11(4): 116–29.

15. Smrekar F. GMP phage production for clinical trials. Ljubljana, Slovenia: AmpliPhi Biosciences; 2014.

16. Authorities NAoT. OECD Principles of Good Laboratory Practice and NATA GLP Compliance Monitoring Program. Sydney: NATA, 2026.

17. Pirnay JP, Blasdel BG, Bretaudeau L, et al. Quality and safety requirements for sustainable phage therapy products. Pharm Res 2015; 32(7): 2173–9.

18. Hibstu Z, Belew H, Akelew Y, Mengist HM. Phage Therapy: A Different Approach to Fight Bacterial Infections. Biologics 2022; 16: 173–86.

19. Pirnay JP, Verbeken G, Ceyssens PJ, et al. The Magistral Phage. Viruses 2018; 10(2).

20. Pirnay JP, Verbeken G. Magistral Phage Preparations: Is This the Model for Everyone? Clin Infect Dis 2023; 77(Suppl 5): S360–S9.

21. Pharmaceutical Inspection Co-operation S. PIC/S Guide to Good Manufacturing Practice for Medicinal Products: Part I (PE 009-17). Geneva, Switzerland: PIC/S Secretariat, 2023.

22. Pharmaceutical Inspection Co-operation S. PIC/S Guide to Good Practices for the Preparation of Medicinal Products in Healthcare Establishments (PE 010-4). Geneva, Switzerland: PIC/S Secretariat, 2014.

23. Australia S. Guide to AS/NZS ISO 9001 for quality management system requirements: key guidelines and benefits. Sydney: Standards Australia, 2022.

24. Faltus T. The Medicinal Phage-Regulatory Roadmap for Phage Therapy under EU Pharmaceutical Legislation. Viruses 2024; 16(3).

25. Scheme PICo. PIC/S Guide to Good Manufacturing Practice for Medicinal Products. Annex 1: Manufacture of Sterile Medicinal Product. Geneva: Pharmaceutical Inspection Co-operation Scheme, 2022.

26. Alliance AHI. Australasian Health Facility Guidelines: Part B – Health Facility Briefing and Planning, 0560 Pharmacy Unit. Sydney, Australia: Australasian Health Infrastructure Alliance, 2026.

27. Commission BP. Supplementary Chapter SC I C1. Annex: Guidelines for using the Test for Bacterial Contamination. British Pharmacopoeia 2026. 2026 ed. London: British Pharmacopoeia Commission; 2026.

28. Bretaudeau L, Tremblais K, Aubrit F, Meichenin M, Arnaud I. Good Manufacturing Practice (GMP) Compliance for Phage Therapy Medicinal Products. Front Microbiol 2020; 11: 1161.

29. Tanir T, Orellana M, Escalante A, Moraes de Souza C, Koeris MS. Manufacturing Bacteriophages (Part 1 of 2): Cell Line Development, Upstream, and Downstream Considerations. Pharmaceuticals (Basel) 2021; 14(9).

30. Agency EM. Note for guidance on minimising the risk of transmitting animal spongiform encephalopathy agents via human and veterinary medicinal products (EMA/410/01 Rev.3). London: EMA, 2011.

31. Higashiyama K, Yuan Y, Hashiba N, Masumi-Koizumi K, Yusa K, Uchida K. Quantitation of Residual Host Cell DNA in Recombinant Adeno-Associated Virus Using Droplet Digital Polymerase Chain Reaction. Hum Gene Ther 2023; 34(11-12): 578–85.

32. Organisation WH. WHO Technical Report Series No. 978. Annex 3: Evaluation of animal cell cultures and characterisation of cell banks (includes residual host-cell DNA guidance). Geneva: World Health Organisation, 2013.

33. Fuerst-Wilmes M, Respondek V, Schramm M, Lilienthal N, Buss K, Duechting A. Regulation of phage therapy medicinal products: developments, challenges, and opportunities. Front Cell Infect Microbiol 2025; 15: 1631359.

34. Use ICfHoTRfPfH. ICH Q6B: Specifications – test procedures and acceptance criteria for biotechnological/biological products. Geneva: ICH, 1999.

35. Moon K, Coxon C, Ardal C, et al. Considerations and perspectives on phage therapy from the transatlantic taskforce on antimicrobial resistance. Nat Commun 2025; 16(1): 10883.

36. El-Sayed D, Elsayed T, Amin N, Al-Shahaby A, Goda H. Evaluating the Phenotypic and Genomic Characterisation of Some Egyptian Phages Infecting Shiga Toxin-Producing Escherichia coli O157:H7 for the Prospective Application in Food Bio-Preservation. Biology (Basel) 2022; 11(8).

37. Administration TG. Australian Regulatory Guidelines for Biologicals (ARGB). Appendix 5: Guidance on TGO 85. Canberra: Therapeutic Goods Administration, 2021.

38. Agency EM. Guideline on the sterilisation of the medicinal product, active substance, excipient and primary container. London: European Medicines Agency, 2019.

39. Convention USP. 788 Subvisible particulate matter in injections. United States Pharmacopeia and National Formulary (USP–NF) 2026. Rockville, MD: United States Pharmacopeial Convention; 2026.

40. HealthCare EDftQoM. European Pharmacopoeia policy on bacterial endotoxins in substances for pharmaceutical use. Strasbourg: EDQM, 2026.

41. Chung KM, Nang SC, Tang SS. The Safety of Bacteriophages in Treatment of Diseases Caused by Multidrug-Resistant Bacteria. Pharmaceuticals (Basel) 2023; 16(10).

42. Convention USP. 85 Bacterial endotoxins test. United States Pharmacopeia and National Formulary (USP–NF) 2026. 2026 ed. Rockville, MD: United States Pharmacopeial Convention; 2026.

43. Commission EP. Adoption of revised general monographs 2034 and 2619: inclusion of N-nitrosamine control requirements. Strasbourg: European Directorate for the Quality of Medicines & HealthCare (EDQM), 2026.

44. HealthCare EDftQoM. 2.5.42 N-nitrosamines in active substances. European Pharmacopoeia 116. 11.6 ed. Strasbourg: EDQM; 2026.

45. Administration TG. Microbiological quality of prescription and over-the-counter medicines: Guidance for sponsors. Canberra: Therapeutic Goods Administration, 2018.

46. HealthCare EDftQoM. Microbiological quality of non-sterile pharmaceutical preparations and substances for pharmaceutical use. European Pharmacopoeia 116. 11.6 ed. Strasbourg: EDQM; 2026.

47. Standardization IOf. ISO 10705-3:2024. Water quality – Detection and enumeration of bacteriophages – Part 3: Validation of methods for concentration of bacteriophages from water. Geneva: ISO; 2024.

48. Standardization IOf. ISO 15189:2022. Medical laboratories – Requirements for quality and competence. Geneva: ISO; 2022.

49. Anderson B, Rashid MH, Carter C, et al. Enumeration of bacteriophage particles: Comparative analysis of the traditional plaque assay and real-time QPCR- and nanosight-based assays. Bacteriophage 2011; 1(2): 86–93.

50. Wiebe KG, Cook BWM, Lightly TJ, Court DA, Theriault SS. Investigation into scalable and efficient enterotoxigenic Escherichia coli bacteriophage production. Sci Rep 2024; 14(1): 3618.

51. Organisation WH. Building the evidence for the use of bacteriophage therapy. Geneva: World Health Organisation, 2023.

52. Richter L, Ksiezarczyk K, Paszkowska K, et al. Adsorption of bacteriophages on polypropylene labware affects the reproducibility of phage research. Sci Rep 2021; 11(1): 7387.

53. Huang W, Khan Mirzaei M, Deng L. Comparative evaluation of long-term preservation methods for morphologically distinct bacteriophages. Microbiol Spectr 2025; 13(7): e0144224.

54. Wang Y, He GX, Chiang L, Loeb SK. Evaluating the solar inactivation of enveloped and non-enveloped bacteriophage using biological weighting functions in natural and simulated waters. Water Research 2026; 289.

55. Chan HK, Chang RYK. Inhaled Delivery of Anti-Pseudomonal Phages to Tackle Respiratory Infections Caused by Superbugs. J Aerosol Med Pulm Drug Deliv 2022; 35(2): 73–82.

56. Guglielmotti DM, Mercanti DJ, Reinheimer JA, Quiberoni Adel L. Review: efficiency of physical and chemical treatments on the inactivation of dairy bacteriophages. Front Microbiol 2011; 2: 282.

57. Association IAT. Dangerous Goods Regulations. 65th ed. Montreal: International Air Transport Association; 2024.

1. Haifer C, Kelly CR, Paramsothy S, et al. Australian consensus statements for the regulation, production and use of faecal microbiota transplantation in clinical practice. Gut 2020; 69(5): 801–10.

2. Pirnay JP, Verbeken G, Ceyssens PJ, et al. The Magistral Phage. Viruses 2018; 10(2).

3. Commission EMAHoMAE. Joint statement on the Hospital Exemption for advanced therapy medicinal products. Amsterdam: EMA, 2025.

4. Moon K, Coxon C, Ardal C, et al. Considerations and perspectives on phage therapy from the transatlantic taskforce on antimicrobial resistance. Nat Commun 2025; 16(1): 10883.

5. Procaccia C. Phage therapy: the medicine of yesterday and tomorrow. Paris: Office Parlementaire d’Évaluation des Choix Scientifiques et Technologiques (OPECST), 2021.

6. Fauconnier A. Phage Therapy Regulation: From Night to Dawn. Viruses 2019; 11(4).

7. Fuerst-Wilmes M, Respondek V, Schramm M, Lilienthal N, Buss K, Duechting A. Regulation of phage therapy medicinal products: developments, challenges, and opportunities. Front Cell Infect Microbiol 2025; 15: 1631359.

9. Suleman M, Clark JR, Bull S, Jones JD. Ethical argument for establishing good manufacturing practice for phage therapy in the UK. J Med Ethics 2025; 51(5).

10. Agency MaHpR. Multi Agency Regulatory Groundwork for Engineered Phage: Regulatory Sandbox Framewor. London: MHRA, 2025.

11. Fukaya-Shiba A, Ogata A, Kuribayashi R, et al. Regulatory considerations for developing phage therapy medicinal products for the treatment of antimicrobial resistant bacterial infections. Front Pharmacol 2025; 16: 1713471.

12. US Food and Drug Administration CfBEaR. Science and regulation of bacteriophage therapy. Silver Spring, MD: FDA, 2021.

13. Lu J, Xu L, Wei W, He W. Advanced therapy medicinal products in China: Regulation and development. MedComm (2020) 2023; 4(3): e251.

14. Yang Q, Le S, Zhu T, Wu N. Regulations of phage therapy across the world. Front Microbiol 2023; 14: 1250848.

15. Zaczek M, Gorski A, Weber-Dabrowska B, et al. A Thorough Synthesis of Phage Therapy Unit Activity in Poland-Its History, Milestones and International Recognition. Viruses 2022; 14(6).

16. Vlassov VV, Tikunova NV, Morozova VV. Bacteriophages as Therapeutic Preparations: What Restricts Their Application in Medicine. Biochemistry (Mosc) 2020; 85(11): 1350–61.

17. Khatami A, Foley DA, Warner MS, et al. Standardised treatment and monitoring protocol to assess safety and tolerability of bacteriophage therapy for adult and paediatric patients (STAMP study): protocol for an open-label, single-arm trial. BMJ Open 2022; 12(12): e065401.

18. Khatami A, Lin RCY, Petrovic-Fabijan A, et al. Bacterial lysis, autophagy and innate immune responses during adjunctive phage therapy in a child. EMBO Mol Med 2021; 13(9): e13936.

19. Petrovic Fabijan A, Lin RCY, Ho J, et al. Safety of bacteriophage therapy in severe Staphylococcus aureus infection. Nat Microbiol 2020; 5(3): 465–72.

20. Administration TG. Proposed GMP exemption for certain bacteriophage manufacture: consultation paper. Canberra: TGA, 2026.

21. Chung KM, Nang SC, Tang SS. The Safety of Bacteriophages in Treatment of Diseases Caused by Multidrug-Resistant Bacteria. Pharmaceuticals (Basel) 2023; 16(10).

22. Commission BP. Supplementary Chapter SC I C1. Annex: Guidelines for using the Test for Bacterial Contamination. British Pharmacopoeia 2026. 2026 ed. London: British Pharmacopoeia Commission; 2026.

